# Evaluating the yaws diagnostic gap: a survey to determine the capacity of and barriers to improving diagnostics in all yaws-endemic countries

**DOI:** 10.1101/2022.05.30.22275669

**Authors:** Becca L. Handley, Serges Tchatchouang, Lise Grout, Roch Christian Johnson, Earnest Njih Tabah, Bernard Boua, Alphonse Um Boock, Aboa Paul Koffi, Delphin Mavinga Phanzu, Nana Konama Kotey, Emerson Rogers, Belen Dofitas, Younghee Jung, Tchalim Maweke, Camila G. Beiras, Issaka Maman, Laud Anthony Basing, Solange Ngazoa, Jean Gabin Houezo, Kwamy Togbey, Elizabeth Freda Telan, Nevio Sarmento, Estelle Marion, Kwasi Kennedy Addo, Oriol Mitjà, Kinsley Asideu, Emma Harding-Esch, Michael Marks

## Abstract

**Background:** Yaws, caused by *Treponema pallidum* subsp. *pertenue*, is a skin neglected tropical disease. It is targeted for eradication by 2030, primarily using mass drug administration (MDA) with azithromycin. Traditionally, diagnosis of yaws has relied on clinical examination and serological testing. However, these approaches have poor diagnostic performance. To achieve eradication, more accurate diagnostics are required to determine whether MDA should be initiated or continued as well as for post-elimination surveillance. Molecular tools will be crucial for detecting antimicrobial resistant cases, which have the potential to derail eradication efforts. In order to determine the feasibility of introducing novel, more accurate, diagnostics for yaws surveillance purposes, it is necessary to understand current in-country diagnostic capacity. This study therefore aimed to understand the current capacity of, and challenges to, improving diagnostics for yaws in all yaws-endemic countries worldwide.

**Methodology/ principal findings:** An online survey was sent to all 15 yaws-endemic countries in July 2021. The survey asked about past prevalence estimates, the availability of different diagnostic tools, and perceived barriers to enhancing capacity. Fourteen countries responded to the survey, four of which did not have a current National Policy for yaws eradication in place. Over 95% of reported that yaws cases from the past five years had not been confirmed with serological or molecular tools, largely due to the limited supply of rapid serological tests. Only four countries reported having operational laboratories for molecular yaws diagnosis, with only one of these having a validated assay to detect azithromycin resistance.

**Conclusions and Significance:** This study highlights the diagnostic capacity constraints across all respondent countries. Countries are in need of access to a sustainable supply of serological tests, and development of molecular testing facilities. Sufficient sustainable funding should be made available to ensure that appropriate diagnostic tools are available and utilised.

## Introduction

Yaws is a neglected tropical disease (NTD) targeted for eradication by 2030^1^. The disease, caused by the spirochete bacteria *Treponema pallidum* subspecies *pertenue (TPE)*, primarily infects children aged between 5-15, living in endemic areas of the tropics^2–4^. It is spread through skin-to-skin contact and characterised by distinct clinical stages interspersed with periods of latency, where the person is asymptomatic. If left untreated, a person may develop the non-infectious but destructive tertiary stage of the disease, often 5-10 years after the initial infection^4,5^.

Yaws was once a global disease, affecting 160 million people in 88 countries and was considered a major public health problem due to the development of deformative and destructive infections occurring on a massive scale^6^. However, a World Health Organization (WHO)/ United Nations Children’s Fund (UNICEF)-run eradication campaign, which took place during the 1950’s and 60’s, reduced worldwide prevalence by up to 95%^7^. This was primarily achieved through the use of mass treatment of injectable benzathine penicillin to yaws patients and their close contacts^8^. Despite the success of the campaign, small clusters of cases remained and have rebounded in recent years^9;^; as of 2021, yaws was confirmed to still be endemic in at least 15 countries^10^. In 2012, after the demonstration that oral azithromycin was as effective as penicillin at treating yaws^11^, a new eradication effort was launched by WHO, known as the Morges strategy^1^. Initially yaws was targeted for eradication by 2020, but this has been extended to 2030 in the new NTD roadmap^12^. The Morges strategy involves the identification and mass drug administration (MDA) of yaws-endemic communities, known as total community treatment (TCT), followed by targeted treatment of any cases that remain and their close contacts, known as total targeted treatment (TTT)^1^. In order for a country to be certified by WHO as having achieved elimination, three criteria must be met: 1) the absence of new serologically confirmed indigenous cases for three consecutive years; 2) the absence of any PCR confirmed cases; 3) the absence of evidence of transmission for three continuous years, which should be measured with sero-surveys among children aged 1–5 years^13^.

Yaws is often diagnosed based on clinical presentation alone. However, this approach has poor diagnostic performance as many clinically-suspected yaws cases are caused by other pathogens and would therefore be negative by serological testing^2^. WHO recommends using sequential testing of two serological tests, the first to detect long-lived treponemal antibodies, followed by a DPP® Syphilis Screen & Confirm Assay (hereafter DPP) (Chembio, Medford, New York, United Kingdom) to confirm current yaws infection ^14^. This is done as the latter test is more expensive, and so is only used when exposure (past or current) to a treponeme is confirmed. However, even when a patient presents with reactive serology (dual positive on a DPP test), only around one-third of people will be positive for TPE DNA by PCR, confirming the current lesion is due to yaws^15–17^. Many patients with clinically-suspected yaws (up to 50% in some settings) are found to harbour *Haemophilus ducreyi* and are negative for TPE bacteria, and around a third of yaws-like lesions are negative for both bacteria and have an unknown aetiology ^15,16,18,19^. Because of the diagnostic accuracy challenges of serology, molecular tools are the preferred diagnostic method used by researchers and yield the most specific diagnostic results. Molecular tools are also the only available diagnostic tool for detecting azithromycin resistant TPE ^20,21^. This is important, as although only recently detected in TPE, drug resistance has the potential to derail eradication efforts ^22,23^.The latest WHO manual on yaws eradication for programme managers^24^, highlights the importance of molecular tools both for declaring interruption of yaws transmission and for surveillance after eradication certification. However, molecular tools are expensive, require trained technicians to carry out the tests and are often only available at larger, national laboratories that are far from the patient.

Despite the eradication target being less than a decade away, the majority of yaws cases reported to WHO are suspected cases, that is, people diagnosed based on clinical examination only. According to data from the Global Health Observatory, only 153 of the 82,564 reported suspected yaws cases were serologically confirmed using a DPP test in 2020 ^10^. Although it is unclear how many suspected cases were tested with any form of serological tests, the low number of serologically confirmed cases indicates there is a major shortfall in the use of appropriate diagnostic techniques. The 2021-2030 NTD roadmap document lays out the desired goals for all NTDs and proposes concrete plans to aid with reaching these targets^12^, including a gap assessment covering diagnostics for each disease. Overall, yaws diagnostics are classed as “green”, indicating diagnostics in their current form are unlikely to impede meeting the eradication target. However, the specific assessment of diagnostic gaps and priorities is classed as “yellow”, with the need to develop both a sensitive molecular point of care (POC) test that can distinguish between TPE and *H. ducreyi* and tools to detect azithromycin resistance.

Thus, if the eradication programme is to be successful, programmatic access to sensitive and specific diagnostic tools will be vital. It is essential that the prevalence of yaws is well mapped initially to determine where eradication efforts should be targeted. It is also key that sensitive diagnostic methods, such as PCR, are routinely available post-TCT to make decisions about continuing eradication efforts or to enable certification of eradication. There is also a clear need for tools that can diagnose azithromycin-resistant TPE. Based on WHO reporting data, we suspect that most endemic countries do not have access to adequate diagnostic tools that will be required for eradication. To determine the current status and availability of yaws diagnostics, we worked with national programmes of yaws-endemic countries to compile data on current diagnostic capacity for yaws and to indicate where diagnostic gaps remain.

## Methods

A two-part survey (supplementary material A) was created and provided to the yaws or NTD programme managers in all 15 known yaws-endemic countries: Benin, Cameroon, Central African Republic, Côte d’Ivoire, Congo, Democratic Republic of Congo, Ghana, Indonesia, Liberia, Papua New Guinea, Philippines, Solomon Islands, Timor-Leste, Togo and Vanuatu. The survey was available in French and English and was available both as an online survey and a paper-based questionnaire.

Reponses were collected between July 2021 – January 2022. If the respondent preferred, an online meeting was arranged and the questionnaire completed by BH, (ST for francophone countries) and the respondents. This exercise was considered an evaluation of routine programmatic capacity not requiring ethics approval.

The first part of the form was designed to be completed by the yaws/NTD programme manager in each country. It included questions about yaws case reporting and availability of POC serological tests as well as about perceived barriers to improving yaws diagnostics. The second part of the form was designed to be completed by a representative of the national yaws molecular testing facility, if applicable. This part of the survey was designed to determine what molecular tests, if any, were in use for routine yaws diagnostics, current capacity to diagnose azithromycin resistance, and barriers in place to enhancing capacity. The questionnaire additionally asked programmes to identify perceived barriers to wider access to yaws diagnostics.

## Results

The programme manager survey was completed by a representative from 14 yaws-endemic countries. Of these, 12 also completed the laboratory survey. Democratic Republic of Congo and Central African Republic did not complete this laboratory component of the survey as they reported having no access to laboratory facilities for yaws diagnosis.

### Yaws policy

Nine countries currently have a national policy for yaws eradication in place, with Papua New Guinea currently developing one. There is no national policy in place in Benin, Cote d’Ivoire, Central African Republic, or Philippines. Yaws is a notifiable disease in 10 (67%) countries: in Central African Republic, Democratic Republic of Congo, Congo and Philippines there are no legal requirements to report yaws infections to health ministries.

### Yaws case estimates and epidemiology

The percentage of districts with serologically-confirmed cases of yaws ranged from 0-38% (Table 1), with 11/14 countries having at least one district with suspected cases thus requiring confirmation of endemicity status. Solomon Islands had the largest proportion of districts reporting clinically suspected cases; all of its ten health districts base endemicity status on suspected cases only and do not confirm with any other tool, although serologically-confirmed cases have been found in non-programmatic research studies in three districts ^25–27^ and serological screening in all provinces has since been initiated. Cameroon had the largest number of suspected-endemic districts, with 152 of its 190 districts having an unconfirmed suspected endemicity status.

**Table 1:** A breakdown of suspected, treponemal rapid diagnostic test (RDT)-confirmed, and serologically confirmed (DPP positive) health districts for each respondent country up to 2021. Here all the categories are exclusive, meaning that each district is listed only once, under its highest level of diagnostic category i.e. a district reporting suspected cases, treponemal RDT positive and DPP positive cases, would only be listed once under “Total number of districts reporting DPP positive cases.”

| Country | Total number of health districts* in the country | Total number of districts reporting DPP positive cases (%) | Percentage of districts reporting DPP positive cases | Total number of health districts reporting treponemal-RDT positive cases | Percentage of health districts reporting treponemal-RDT positive cases | Number of districts reporting clinically-suspected cases | Percentage of districts reporting clinically-suspected cases |
| --- | --- | --- | --- | --- | --- | --- | --- |
| BEN | 77 | 2 | 2.6 | 1 | 1.3 | 4 | 5.2 |
| CAF | 35 | 3 | 8.6 | 2 | 5.7 | 4 | 11.4 |
| CIV | 113 | 15 | 13.3 | Unknown | NA | 98 | 86.7 |
| CMR | 190 | 38 | 20 | 0 | 0 | 152 | 80 |
| COG | 52 | 4 | 7.7 | 0 | 0 | 10 | 19.2 |
| DRC | 516 | 3 | 0.6 | 5 | 1 | 10 | 1.9 |
| GHA | 260 | 21 | 8.1 | 40 | 15.4 | 93 | 35.8 |
| LBR | 93 | 3 | 3.2 | 0 | 0 | 3 | 3.2 |
| PNG | 89 | 0 | 0 | 0 | 0 | 75 | 84 |
| PHL | 243 | 3 | 1.2 | 6 | 2.5 | Unknown | NA |
| SLB | 10 | 0 | 0 | 0 | 0 | 10 | 100 |
| TGO | 39 | 3 | 7.7 | Unknown | NA | Unknown | NA |
| TLS | 13 | 2 | 15.3 | 4 | 30.8 | 0 | 0 |
| VUT | 6 | 5 | 83.3 | 0 | 0 | 1 | 16.7 |

\* The data has been provided for provinces in SLB and VUT
\*\*not confirmed using serological tests.
DPP: DPP® Syphilis Screen & Confirm Assay; RDT: Rapid Diagnostic Test; BEN: Benin; CAF: Central African Republic; CIV: Côte d'Ivoire; CMR: Cameroon; COG: Congo; DRC: Democratic Republic of Congo; GHA: Ghana; LBR: Liberia; PNG: Papua New Guinea; PHL: Philippines; SLB: Solomon Islands; TGO: Togo; TLS: Timor-Leste; VUT: Vanuatu.

Respondents were asked for a breakdown of their national yaws reporting data from the last two years that data were reported to WHO since 2016, including the number of suspected cases, tests performed and number of positive results. No data were reported to WHO from Central African Republic over the past five years. Data from the last two years of reporting, 2019 and 2020, are displayed in Figure 1A. In 2020, nine countries shared data with WHO, with the number of suspected cases ranging from 121 in Timor-Leste, to 81,369 cases in Papua New Guinea. Ghana and Timor-Leste reported that they tested all suspected cases with a treponemal antibody test, whereas Papua New Guinea and Solomon Islands reported based solely on clinical diagnosis. Despite recording over 10,000 suspected cases in Cote d’Ivoire of which 30% were tested using treponemal RDTs and 74% of positive RDTs were tested with DPP tests, only 26 cases of serologically confirmed yaws cases were detected in 2020.

**Figure 01:**
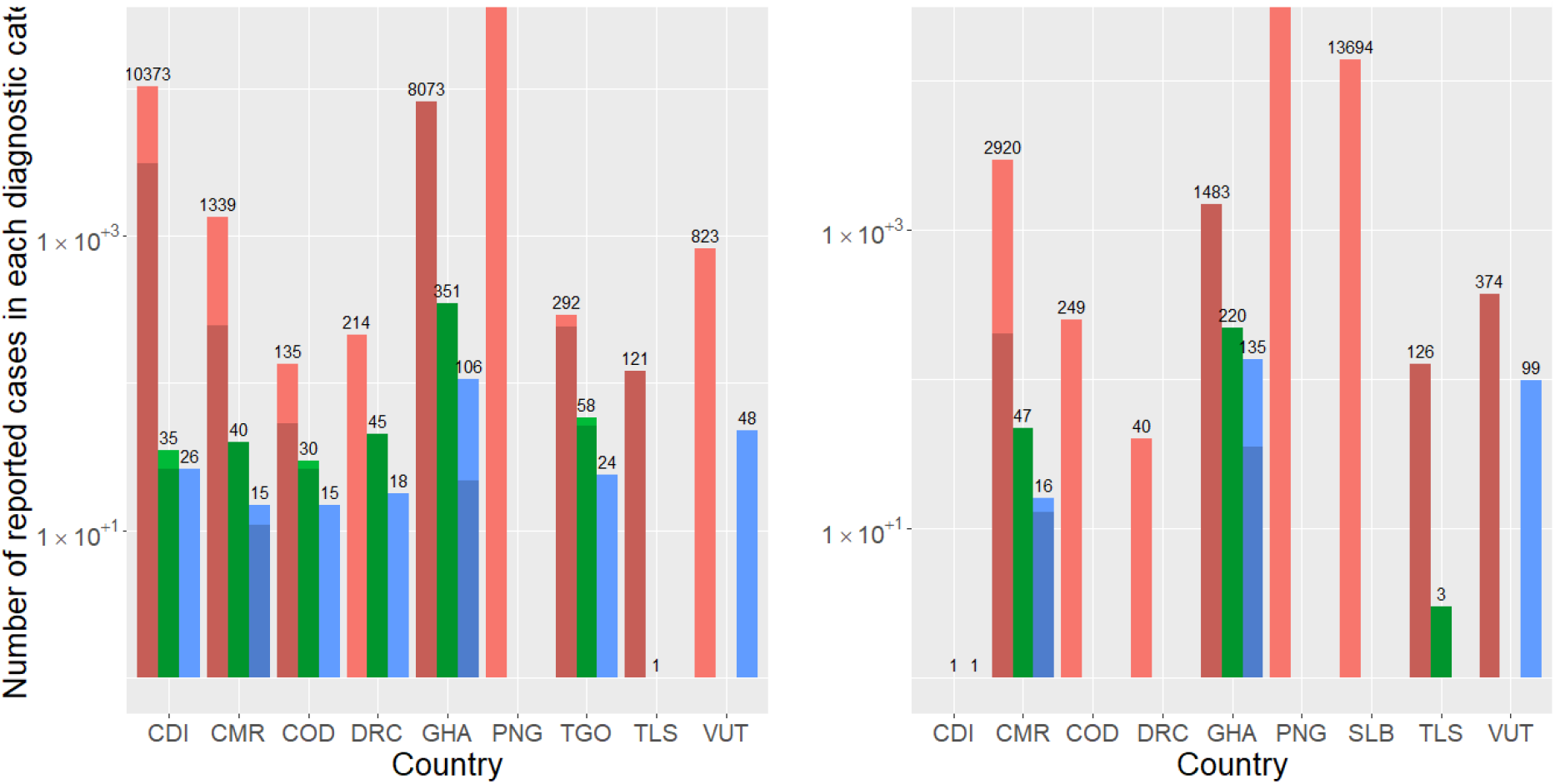
The number of suspected, treponemal rapid diagnostic test positive (RDT), dual path platform syphilis screen (DPP) positive cases reported by each country in (A) 2020 and (B) 2019, plotted on logarithmic scale. Shaded areas of the bars represent the number of people tested using sequential testing (treponemal-RDT, DPP, PCR). Categories are not exclusive as a person can fall under multiple categories.

In 2019, nine countries reported data to WHO (Fig. 1B), with four countries (Congo, Côte d’Ivoire, Papua New Guinea and Solomon Islands) reporting suspected cases only. However, two countries (Ghana, Timor-Leste) reported performing sequential serological testing (treponemal-RDT and DPP) on all positive patients and Vanuatu used DPP testing on all suspected yaws cases. Only Cameroon and Ghana performed PCR tests on DPP positive cases. Cameroon reported no positive PCR cases either year and Ghana reported two and one positive case from 2019 and 2020, respectively.

### Diagnostic capacity of health care workers

In four countries (Benin, Côte d’Ivoire, Philippines, Papua New Guinea) there was no standardised training taking place for health care workers to detect yaws, and with no training planned in two of these countries (Benin and Côte d’Ivoire). In countries where training has taken place, this was occasionally combined with training on the recognition of other skin NTDs, such as in Solomon Islands and Vanuatu who implemented integrated skin NTD training in 2020.. All but one respondent country (Papua New Guinea) said their healthcare workers had access to the WHO yaws recognition booklet for communities^20^; however, these were not available in all health districts that required them. In 11 countries, health care workers trained to conduct treponemal-RDTs and DPP tests were not available in all suspected- or known-endemic districts.

### Serological tools

Figure 2 shows which countries routinely use which serological tests (treponemal-RDTs or DPPs). Six of the 14 countries advise to routinely test all suspected yaws cases with treponemal RDTs, but four of these say the tests are not always available for patients presenting at clinics (Côte d’Ivoire, Ghana, Liberia or Togo) and Liberia reported tests are also not always available during routine surveillance activities. In Vanuatu, the guidelines state that all suspected cases should be tested with a DPP only, whereas Central African Republic, Côte d’Ivoire, Ghana, Liberia, Timor-Leste, and Togo use DPPs according to the suggested WHO sequential testing strategy (only on patients with a positive treponemal RDT test). Again, DPPs are not routinely available to test all patients with suspected yaws presenting at health care centres (Côte d’Ivoire, Ghana, Liberia, Timor-Leste or Vanuatu) or during routine case finding (Liberia, Timor-Leste or Vanuatu).

**Figure 02:**
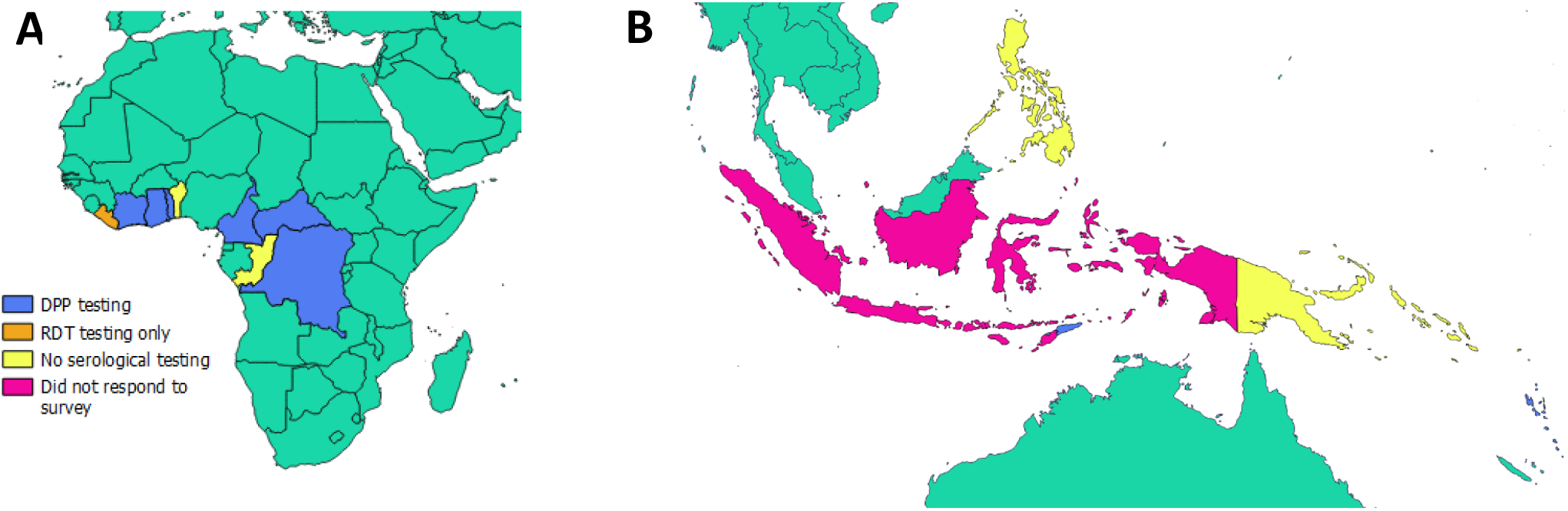
Serological testing strategy of yaws of all yaws endemic countries. (A) Africa, (B) Asia and the Western Pacific

### Molecular tools

Only four countries (Cameroon, Côte d’Ivoire, Ghana, and Togo) reported currently running PCR testing for yaws (Fig.3). No standardised PCR assays were being used across these four countries, however all reported to test for the Pol A pan-treponemal target. There have been PCR confirmed cases in Liberia ^28^, Papua New Guinea^11,29,30^, Solomon Islands^15,26^, Vanuatu^19,31^ through research studies, but the samples were tested abroad. All four countries with laboratory capacity reported having the correct equipment to run the tests and adequate trained staff available. Other countries responded that they had laboratory facilities and equipment in place, but did not have standard operating procedures, trained staff or the necessary reagents and consumables to run the tests.

**Figure 03:**
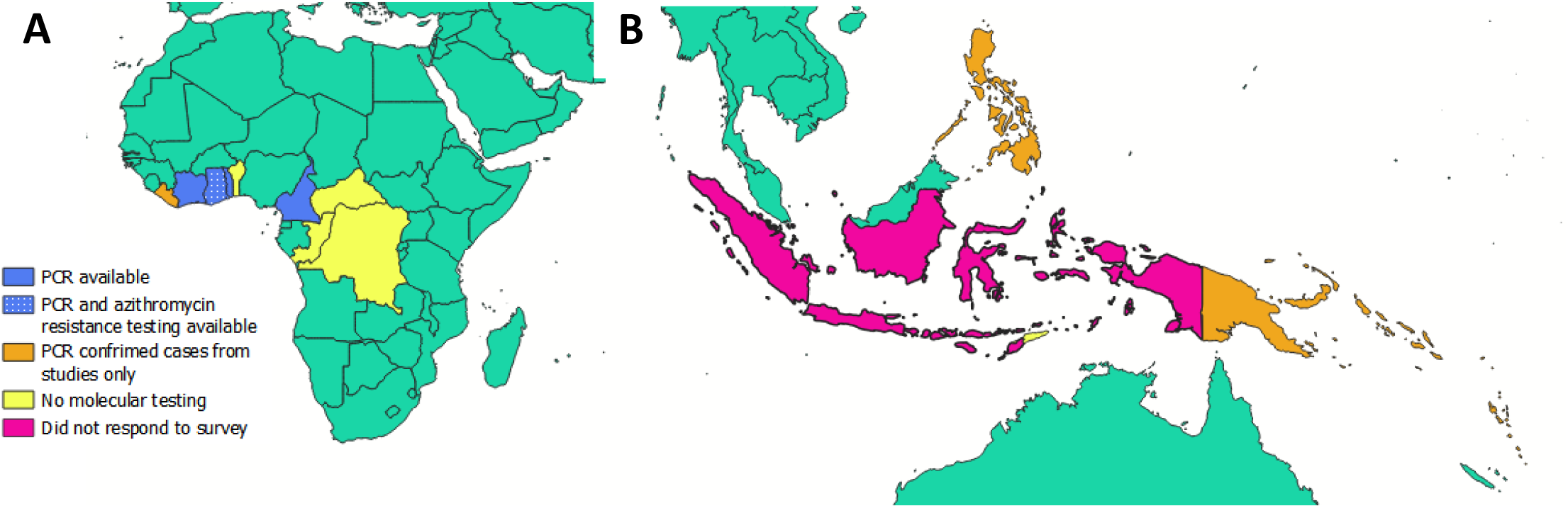
Molecular testing capacity for yaws of all yaws endemic countries. (A) Africa, (B) Asia and the Western Pacific

### Patient follow up and azithromycin resistance detection

In general, countries included routine follow up of azithromycin treated cases to check for treatment failure, in their national guidelines. In Papua New Guinea, Solomon Islands and Vanuatu this was not the case, however none of these countries had started MDA with azithromycin outside of study settings. Of the ten countries that reported routine follow up of patients, six (Benin, Côte d’Ivoire, Democratic Republic of Congo, Ghana, Timor-Leste, and Togo) stated that 100% of those treated are followed up. In Central African Republic and Congo, it was unknown how many patients were re-visited, and Cameroon estimated that around 50% of those treated were successfully re-visited within the suggested 2-4 week time frame.

Despite the high follow up rate reported in many countries, swabs were not routinely collected from patients with treatment failure, with only Cameroon and Liberia reporting the routine collection of swabs. In Liberia, these data were limited to a single academic study ^28^. The respondents from Benin, Central African Republic and Togo were unsure if swabs were collected during follow up visits. Only Ghana currently has capacity in-country to perform a quantitative (q)PCR for the detection of azithromycin resistance. Cameroon stated they have established a qPCR assay, however it has not yet been clinically validated. Overall, the ability to promptly detect azithromycin resistance is lacking in 13 (93%) of the respondent yaws-endemic countries.

### Barriers to enhancing diagnostic capacity

In terms of identifying suspected yaws cases, the respondent from the Philippines noted that as there is no established yaws programme, there is no funding available for any yaws-related activities and yaws-endemic local government units have limited funds to support case detection and treatment activities. The lack of trained staff was mentioned by five countries as one of the main barriers to identifying all suspected cases. Lack of funding, including funding to train staff, was specifically mentioned by eight respondents. Other barriers included a lack of community awareness, and difficult-to-access communities and populations. Congo said that they had received project-specific funding to map four known-endemic districts, but this funding did not stretch to the additional ten suspected-endemic districts not involved in the research partnership.

With regards to testing all suspected cases with a treponemal RDT, and confirming all treponemal RDT positive cases with a DPP test, the main barriers were stock outs, lack of sustainable supply and complete absence of the serological tests in-country. Eight countries specifically mentioned there was a lack of funding to purchase the necessary tests. There is a reliance on WHO, programme partners, non-governmental organisations or academic studies to provide tests, with some countries saying they had surplus tests from previous projects, but once these were used there were none available and it was unclear how to acquire more. Most respondents were unsure about the availability of RDTs and DPPs for syphilis, so it remains unclear if there are similar supply issues for detecting the sexually transmitted *T. pallidum* infection. Other identified barriers included a lack of public awareness or difficulties in accessing health care centres.

In relation to improving molecular testing capacity, in Cameroon and Togo, where molecular facilities are available, both reported transport of samples from patient to the reference lab was a barrier to enhancing molecular capacity. Côte d’Ivoire cited a lack of facilities close to the patient as a major obstacle. Ghana reported funding and logistics were the biggest barrier to enhancing molecular testing capacity, including insufficient funding to set up labs in yaws-endemic health districts. In the countries without molecular diagnosis for yaws, funding was again overwhelmingly cited as the main barrier to enhancing capacity. Although many countries have a lab facility that could be utilised for yaws diagnostics, a lack of specific funding means it is not possible to pay for staff, reagents or consumables. Some countries’ respondents suggested there was a lack of awareness of the need for molecular testing, or said there was not a need for these facilities as swabs can be sent to other countries in which molecular testing facilities are already in place.

## Discussion

Whilst the new NTD roadmap outlines that diagnostic testing is intended to play a pivotal role in yaws eradication, the results from this survey demonstrate there is inadequate diagnostic capacity for yaws across nearly all yaws-endemic countries. Improvement in access to current diagnostics, as well as the development of novel molecular diagnostics that can be performed close to the patient and that are able to detect azithromycin resistance, will be key to achieving the 2030 eradication target. Overall, this survey highlighted that funding, including operational and staff funding as well as funding for reagents, and equipment costs were considered the major barrier to improving all forms of diagnostics across most yaws-endemic countries.

One of the most striking findings of this survey was that, despite the eradication target being less than a decade away, five (35%) of 14 respondent countries do not currently have a yaws eradication policy in place, a figure that is essentially unchanged since the 2017 global WHO survey for yaws^32^. In Central African Republic, Congo, Democratic Republic of Congo, and Philippines, yaws is not a notifiable disease, meaning that reported case data are likely to be an underestimate. Successful disease control and eradication programs, such as the eradication of dracunculiasis have relied on having an robust surveillance and reporting system within their national programmes meaning cases are detected and responded to promptly ^33^. Where possible disease surveillance should be integrated amongst NTDs and beyond, to allow for time- and cost-effective disease monitoring^34^.

Our survey demonstrates that the endemicity status of most districts is based on clinical diagnosis rather than serological confirmation and that in 11 countries health care workers were not trained to recognise and diagnose yaws in all suspected districts. An absence of trained healthcare workers creates a major barrier to delivering yaws interventions and surveillance. An unpublished survey of government physicians and dermatologists in the Philippines which aimed to determine if the respondents had ever encountered any yaws cases found only three of the 131 respondents reported seeing yaws cases in the past. Of the remaining respondents the majority were not knowledgeable about yaws ^32^. Work is needed to ensure health care workers in all yaws-endemic countries receive standardised training to recognise yaws and perform appropriate serological testing.

At least two-thirds of countries have are not running any molecular diagnostics for yaws. This contrasts with the NTD roadmap assessment of NTD diagnostics, which classes yaws diagnostics as green and thus not likely to hinder roadmap goals. As countries move towards eradication, molecular testing capacity will become more important as it is the only form of testing that is both sensitive enough to detect single cases of yaws and with an adequate specificity to avoid false positive results and the deployment of unnecessary interventions. Most countries indicated there is infrastructure and staff available to perform this testing if required. Therefore, an achievable priority should be to set up molecular testing capacity in these countries. In order for molecular testing to be carried out sustainably throughout the eradication programme, all countries must also be able to easily purchase all reagents and consumables required to run the tests at a reasonable cost.

Despite being recommended by WHO swabs are not being routinely collected from patients with treatment failure during routine follow up and only Ghana has a validated azithromycin PCR currently in use. They have not yet detected any azithromycin resistance strains, meaning the first line treatment with azithromycin can continue to be used in all endemic areas. In all other yaws endemic countries, there is real potential not only for the emergence of azithromycin resistance, but its uncontrolled spread if not detected promptly. High quality surveillance of antimicrobial resistance is often lacking in low and middle-income countries but enhancing laboratory capacity for detection and surveillance of antimicrobial resistance will be crucial not only for yaws eradication but also to protect against the threat of resistance to other common and life-threatening diseases^35,36^. Overall, many of the above recommendations will only be achievable with sufficient funding made available to national programmes. The new NTD roadmap^12^ rightly outlines a paradigm shift towards country ownership and away from traditional donor-led approaches, thus countries must ensure they have sustainable funding routes in place to acquire and maintain the diagnostic capacity needed for NTDs, including yaws.

There are some limitations to this report. Firstly, this survey was designed for, and only sent to, countries that are currently known to be endemic. Therefore, we do not know what capacity, if any, the 72 previously-endemic countries have to diagnose yaws; these countries may be yaws-free or may simply lack adequate surveillance to detect and report cases. It would be valuable to send a condensed version of this diagnostic gap survey to formerly-endemic current with uncertain status to determine what capacity they may have for the diagnosis of resurgent cases. It is vital that, if there are other endemic countries, health care workers know the clinical signs consistent with yaws and ministries of health are aware of how to act in the event of the detection of a suspected case.

Another limitation of this study is that the majority of countries provided national programme data for this survey, and did not include data from research studies, which can mean health ministries are providing an incomplete picture. For example, in Solomon Islands and Papua New Guinea, the two countries with the highest burden of yaws, there have been no serologically or molecularly confirmed cases of yaws reported to ministries of health; however, due to the inclusion of these two countries in many academic studies ^15,16,26,29,38,39^, it has been demonstrated that a number of districts harbour both serologically and molecularly confirmed cases. Contrastingly, all the data provided here for Liberia is from a single academic study conducted in 2018 ^40^ and no additional programmatic data were provided.

## Conclusions

Yaws diagnostic capacity across all endemic countries is currently lacking. The majority of people with suspected yaws are not being tested with any serological tools, molecular testing is extremely limited and testing for drug resistance almost non-existent. As a result, yaws prevalence is hard to estimate, limiting our ability to prioritise and target eradication efforts. There is a general over-reliance on donors and academic partners for access to diagnostic testing, with many countries struggling with funding for, or access to, serological and molecular tests. Addressing these gaps is critical to achieving the 2030 yaws eradication target.

## Supporting information

A two-part survey (supplementary material A) was created and provided to the yaws or NTD programme managers in all 15 known yaws-endemic countries: Be

A two-part survey (supplementary material A) was created and provided to the yaws or NTD programme managers in all 15 known yaws-endemic countries: Be

## Data Availability

All data produced in the present study are available upon reasonable request to the authors

## Acknowledgments

We are grateful to all the respondents of the survey for taking the time to gather and provide us with the data needed for this manuscript. We are also grateful to Esther Amon for her administrative support during this project.

## Contribution

EMH-E, MM and BLH conceived the idea for the study. BLH drafted the survey with input from LG, KA, EMH-E, MM and ST. BLH and ST conducted interviews. ST translated relevant material. BLH analysed the data and drafted the manuscript. ST, RCJ, ET, BB, EUB, APK, DMP, NKK, ER, BD YJ, Tm, CGB, IM, LAB, SN, JGH, KT, EFT, NS, EM, KKA collated and provided country data. All co-authors read and approved the final draft of this manuscript

