## Supplementary material for "Evaluating the yaws diagnostic gap: a survey to determine the capacity of and barriers to improving diagnostics in all yaws-endemic countries": A two-part survey (supplementary material A) was created and provided to the yaws or NTD programme managers in all 15 known yaws-endemic countries: Be

#### Diagnostic Gap Survey: Part I

#### Please complete the final form online:

#### <https://odk-survey.lshtm.ac.uk/-/single/zqueFvaCxRxRkCmVLzH2QNa1r93jwHf?st=qxCRyyzqt11a4LV$1vBQoAo!eh7GgwDiJavxKZYHgy4ZVEcDdD1J7qNWILqRHJ9Y>

#### Respondent information

#### Name:

#### _________________________________________________

#### Email address:

#### _________________________________________________

#### Country you are representing for this questionnaire:

#### _________________________________________________

#### Your role:

#### _________________________________________________

#### Section 1: Yaws case reporting

1. **Which country are you representing for the purpose of this questionnaire?**

____________________

2. **Is there a national policy for yaws eradication?**

- *Yes*
- *No*
- *Unknown*

3. **Is yaws a notifiable disease?**
   - *Yes*
   - *No*
   - *Unknown*
4. **How many districts are there in your country?**

___________________

5. **In the last five years how many districts in have reported confirmed yaws cases?**

*How many districts have reported serologically confirmed (dual positive) or PCR confirmed cases of yaws? (Input "999" if you are unsure)*

___________________

6. **In the last five years how many districts in have reported treponemal-positive yaws cases?**

*Clinical cases with positive treponemal POC test, positive TPPA or positive TPHA test in districts with no serologically or molecularly confirmed cases of yaws (Input "999" if you are unsure)*

____________________

7. **How many additional health districts are suspected to be endemic for yaws?**

*Reports of clinical cases consistent with yaws but not confirmed using any test. (Input "999" if you are unsure)*

____________________

8. **What district level data are reported to the health ministries in order to generate national yaws figures?**
   - *Suspected cases*
   - *Treponemal-positive cases*
   - *Serologically confirmed cases*
   - *PCR confirmed cases*
   - *Unknown*
9. **What was the most recent year that data was reported to WHO? If data has not been reported in the last five years please leave blank.**

**The next few questions are aimed to find out a breakdown of cases detected/reported to WHO in the most recent year data was reported to WHO*

*___________________*

10. **In the most recent year of data reporting how many cases of suspected yaws were reported? Suspected cases are people living endemic areas presenting with yaws like lesions**

*­*

*_____________________*

11. **Of these patients with suspected yaws how many were tested using a treponemal antibody test?**

_____________________

12. **Of those tested with a treponemal antibody test how many were positive for treponemal antibodies?**

___________________

13. **Of the patients with suspected yaws how many how many were tested using DPP tests OR lab-based treponemal and non-treponemal assays?**

____________________

14. **Of those tested, how many were positive for both treponemal and non-treponemal antibodies and diagnosed with serologically confirmed yaws?**

____________________

15. **Of the patients with suspected yaws how many were tested for yaws using PCR?**

____________________

16. **How many of these were positive by PCR and considered PCR-confirmed yaws cases?**

____________________

17. **Do you have any other information to share about case data collected in the most recent year of data reporting?**

#### ____________________________________________________________

#### ____________________________________________________________

18. **Prior to the most recent year of data reporting when was data most recently reported to WHO? If data has not been reported in the last five years please leave blank.**

_____________________

19. **In the second most recent year how many cases of suspected yaws were reported? Suspected cases are people living endemic areas presenting with yaws like lesions**

__________________________

20. **Of these patients with suspected yaws how many were tested using a treponemal antibody test?**

______________________

21. **Of those tested with a treponemal antibody test how many were positive for treponemal antibodies?**

_____________________

22. **Of the patients with suspected yaws how many how many were tested using DPP tests OR lab-based treponemal and non-treponemal assays?**

____________________

23. **Of those tested, how many were positive for both treponemal and non-treponemal antibodies and diagnosed with serologically confirmed yaws?**

_____________________

24. **Of the patients with suspected yaws how many were tested for yaws using PCR?**

______________________

25. **How many of these were positive by PCR and considered PCR-confirmed yaws cases?**

______________________

26. **Do you have any other information to share about case data collected in the second most recent year of data reporting?**

#### ____________________________________________________________

#### ____________________________________________________________

#### Section 2: Clinical diagnosis

#### *These next few questions are about current standards of clinical diagnosis for yaws*

1. **Is there standardised training taking place for district health workers to recognise yaws?**
   - *Yes*
   - *No*
   - *Unknown*
2. **If no, are there plans for training to occur?**
   - *Yes*
   - *No*
   - *Unknown*
3. **Do healthcare workers have access to WHO yaws training materials?**

*E.g. Yaws: Recognition Booklet for Communities*

- *Yes*
- *No*
- *Unknown*

4. **Do you have any other information about clinical yaws diagnostics that you would like to share?**

________________________________________________________________________________________________________________________________________________________________

#### Section 3: Serological Diagnosis

*These next questions are about the use of serological testing for yaws*

1. **Are healthcare workers in all suspected endemic districts trained to perform treponemal RDTs and DPPs?**
   - *Yes*
   - *No*
   - *Unknown*
2. **Are treponemal RDTs routinely used on patients presenting with clinical yaws? (Tests which only detect treponemal antibodies)**

*Routinely used means there are tests available and it is standard practice to use these on patients with suspected yaws*

- *Yes*
- *No*
- *Unknown*

3. **If q2 = yes, which RDTs are used? (List all that apply)**
4. **Are the treponemal RDTS (whichever are used) routinely available for every suspected yaws case presenting at a health care facility?**

*Routinely available means an RDT is available for every patient with suspected yaws presenting at a health care centre*

- *Yes*
- *No*
- *Unknown*

5. **Are confirmatory tests routinely used (treponemal RDT followed by DPP/RPR/VDRL to detect non-treponemal antibodies)**
   - *Yes*
   - *No*
   - *Unknown*
6. **Are DPPs routinely available for every suspected yaws case presenting at a health care facility?**

*Routinely available means a DPP is available for every patient with suspected yaws presenting at a health care centre*

- *Yes*
- *No*
- *Unknown*

7. **Are the treponemal RDTs routinely available for all patients with suspected yaws found during targeted case finding activities?**
   - *Yes*
   - *No*
   - *Unknown*

*Routinely available means an RDT is available for patients with suspected yaws detected during surveillance activities*

8. **Are DPPs routinely available for all patients with suspected yaws found during targeted case finding activities?**

*Routinely available means a DPP is available for patients with suspected yaws detected during surveillance activities*

- *Yes*
- *No*
- *Unknown*

9. **Do you have any other information you would like to share with us about the serological tests being performed for yaws?**

______________________________________________________________________________________________________________________________________________________________________

#### Section 4: Molecular diagnosis for yaws

#### *These next questions are designed to understand what molecular testing, if any, is currently taking place and what capacity you may have for increasing molecular testing in the future*

#### 1. Has your country got access to molecular testing facilities specifically for yaws?

- *Yes*
- *No*
- *Unknown*

#### 2. If 1 = yes, how many laboratories are currently involved in molecular testing for yaws?

#### _____________________

#### 3. If 1 = yes, where are these laboratory facilities available?

- *National Reference Laboratories*
- *Regional labs*
- *District labs*
- *Sub-district (local)*
- *Unknown*

#### ****If your country has access to molecular testing for yaws please ensure that a representative from the main yaws reference laboratory completes Part II of the Yaws Diagnostic Gap Survey****

#### Section 5: Yaws case management

*The following questions are about yaws patient management*

1. **According to your national yaws guidelines which patients qualify for treatment with azithromycin?**

*select all that apply*

- *Suspected Cases*
- *Probable Cases*
- *Serologically confirmed cases*
- *PCR confirmed cases*
- *Unknown*

2. **Do the national guidelines state you should follow up azithromycin treated patients between 2 and 4 weeks after treatment?**
   - *Yes*
   - *No*
   - *Unknown*
3. **If q2 = yes, what proportion of azithromycin treated patients are followed up?**

*In the last year of data collection, what percentage of people treated with azithromycin for yaws were followed up 2-4 weeks post treatment to detect treatment failure? If unknown type "unknown"*

4. **If q2 = yes, are additional lesion swabs collected from patients with reported treatment failure?**
   - *Yes*
   - *No*
   - *Unknown*
5. **If q4 = yes, from how many patients with yaws treatment failure were swabs collected last year?**

**____________**

**6. Are molecular tests for azithromycin resistance available?**

*Available means that a standard operating procedure is in place in the laboratory, that personnel are trained, and that all reagents and equipment are available to perform these tests upon request.*

- *Yes*
- *No*
- *Unknown*

#### **In-depth questions regarding molecular testing for azithromycin resistance are included in Part II of the Yaws Diagnostic Gaps Survey.

#### Section 6: Syphilis testing capacity

*These questions have been asked to assess what capacity you have for testing syphilis*

1. **Are point of care tests (POCTs) routinely used to confirm a clinical diagnosis of syphilis, for example in patients with genital ulcers?**

*Routinely used means a test is performed on each person presenting with genital ulcers at any given health care centre and that it is in the national guidelines to use these tests*

- *Yes*
- *No*
- *Unknown*

2. **Are the POCTs (whichever are used) available for every person presenting with genital ulcers?**

*Selecting yes means there is a test is available for each person presenting with a genital ulcer at a health care facility. If a health care centre struggles with POC shortages select no.*

- *Yes*
- *No*
- *Unknown*

3. **If you have selected <<yes>> for question two, please list which POCTs are used.**

___________________________________

4. **Are POCTs routinely used for screening pregnant women in antenatal clinics?**

*Routinely used means a test is performed on each pregnant women during her routine antenatal appointments and that it is in the national guidelines to use these tests*

- *Yes*
- *No*
- *Unknown*

5. **Are the POCTs (whichever are used) tests routinely available for every woman being screened at the antenatal clinic?**

*Selecting yes means there test is available for each pregnant women attending her antenatal appointments at a health care facility. If these tests are not readily available at all health care centres involved in antenatal screening select no.*

- *Yes*
- *No*
- *Unknown*

**Section 7: Funding for serological testing**

*These next questions are designed to find out about funding for serological testing*

1. **Is there sufficient funding available to pay for equipment needed for serological testing (e.g. lancets or PPE)? Sufficient means enough funding to cover the total costs**
   - *Yes*
   - *No*
   - *Unknown*
2. **What are the sources of funding for equipment?**

- *Health ministries*
- *Programme partner*
- *Other NGO*
- *WHO*
- *Other*
- *Unknown*

3. **Of those you have selected rank them in order of greatest contributor to funding. If not known move on to the next question. List those selected in order of greatest funder.**

**1.**

**2.**

**3.**

**4.**

**5.**

**6.**

4. **Is there sufficient funding available to pay for all staff costs to perform serological testing? Sufficient means enough funding to cover the total costs.**

- *Yes*
- *No*
- *Unknown*

5. **What are the sources of funding for staff costs?**
   - *Health ministries*
   - *Programme partner*
   - *Other NGO*
   - *WHO*
   - *Other*
   - *Unknown*
6. **Of those you have selected rank them in order of greatest contributor to funding. If not known move on to the next question. List those selected in order of greatest funder.**

**1.**

**2.**

**3.**

**4.**

**5.**

**6.**

7. **Is there sufficient funding available to pay for the required treponemal RDT kits? Sufficient means enough funding to cover the total costs**
   - *Yes*
   - *No*
   - *Unknown*
8. **What are the sources of funding for treponemal RDTs?**
   - *Health ministries*
   - *Programme partner*
   - *Other NGO*
   - *WHO*
   - *Other*
   - *Unknown*
9. **Of those you have selected rank them in order of greatest contributor to funding. If not known move on to the next question.**

**1.**

**2.**

**3.**

**4.**

**5.**

**6.**

10. **Is there sufficient funding available to pay for the required DPP kits? Sufficient means enough funding to cover the total costs**
    - *Yes*
    - *No*
    - *Unknown*
11. **Is there a different source of funding for confirmatory tests (DPPs) compared with treponemal RDTs?**
    - *Yes*
    - *No*
    - *Unknown*
12. **If 11 = yes, what are the sources of funding for DPPs?**
    - *Health ministries*
    - *Programme partner*
    - *Other NGO*
    - *WHO*
    - *Other*
    - *Unknown*
13. **If you have selected alternative sources of funding in q12, of those you have selected rank them in order of greatest contributor to funding. If not known move on to the next question.**

**1.**

**2.**

**3.**

**4.**

**5.**

**6.**

14. **Where is the supply of POCT held?**
    - *National reference laboratories*
    - *Regional health care centres / labs*
    - *District health care centres / lab*
    - *Sub-district (local) health care centres or labs*
15. **Where is the supply of DPP held?**
    - *National reference laboratories*
    - *Regional health care centres / labs*
    - *District health care centres / lab*
    - *Sub-district (local) health care centres or labs*

#### Section 8: Barriers to improving diagnostic capacity

*These final questions are designed to see what you believe are the biggest barriers/obstacles for increasing yaws diagnostic capacity. These barriers could be things like lack of funding, staff availability and/or difficulty accessing resources.*

1. **What barriers do you see to identifying all suspected yaws cases?**
2. **What barriers do you see to testing all suspected yaws cases with a treponemal RDT?**
3. **What barriers do you see to testing all treponemal-confirmed yaws cases with DPP?**
4. **What barriers do you see to enhancing molecular diagnostic capacity?**
