## Supplementary material for "Evaluating the yaws diagnostic gap: a survey to determine the capacity of and barriers to improving diagnostics in all yaws-endemic countries": A two-part survey (supplementary material A) was created and provided to the yaws or NTD programme managers in all 15 known yaws-endemic countries: Be

#### Diagnostic Gap Survey: Part II

#### Please complete the final form online:

#### <https://odk-survey.lshtm.ac.uk/-/single/FBRduIaAWLmgzYifyKsCYyEYsCT7B8L?st=enEyTwodGupHAbdyh!HlgaKht!8$0r0gc!UPtR3Lfq9d4f3yNfZ27l0BqxAiR$E8>

#### Respondent information

#### Name:

#### _________________________________________________

#### Email address:

#### _________________________________________________

#### Country you are representing for this questionnaire:

#### _________________________________________________

#### Laboratory you are representing for this questionnaire:

#### _________________________________________________

#### Section 1: Molecular diagnosis

#### *These questions are designed to understand what molecular testing, if any, is currently taking place and what capacity you may have for increasing molecular testing in the future*

1. **Has your country got access to molecular testing facilities specifically for yaws?**
   - *Yes*
   - *No*
   - *Unknown*
2. **If yes, how many laboratories are currently involved in molecular testing for yaws?**

_______________

3. **Where are these laboratory facilities available?**

(select all that apply)

- *National Reference Laboratories*
- *Regional labs*
- *District labs*
- *Sub-district (local)*
- *Unknown*

4. **Are trained staff available at all labs currently conducting molecular testing for yaws?**
   - *Yes*
   - *No*
   - *Unknown*
5. **Which tests are being deployed in these labs?**

Which PCR targets are being detected?

- *polA*
- *tp0548*
- *tp47*
- *Tprl*
- *Unknown*

6. **Are all reagents and consumables readily available to test every swab sent to the lab?**

*Reagents are all chemicals required to run the test, consumables are all other non-specific equipment required such as gloves and tubes.*

- *Yes*
- *No*
- *Unknown*

7. **Is all necessary equipment (e.g. freezers, thermocyclers) to perform these molecular tests available and working?**
   - *Yes*
   - *No*
   - *Unknown*
8. **Do you have any other information you would like to share with us about molecular diagnostics for yaws?**

***____________________________________________________________________________________________________________________________________________***

#### Section 2: Molecular testing for Azithromycin Resistance

1. **Are additional lesion swabs collected from patients with reported treatment failure?**
   - *Yes*
   - *No*
   - *Unknown*
2. **From how many patients with yaws treatment failure were swabs collected last year?**

_________________

3. **Are there molecular tests available to test for azithromycin resistance?**

*Available means an SOP in place in a the lab, with trained staff and all reagents and equipment available. This does not need to be in place specifically for yaws and could be used for detecting azithromycin resistance syphilis*

- *Yes*
- *No*
- *Unknown*

**4. What molecular tests are used to test for azithromycin resistance?**

__________________________

**5. Are these molecular tests routinely used for yaws treatment failure?**

- *Yes*
- *No*
- *Unknown*

**6. How many patients were tested for azithromycin resistant *T. pallidum* in the last year of data collection?**

________________________

**7. How many patients were tested for azithromycin resistant *T. pallidum* in the previous year?**

**­­­­­­­­­­­­­­**

_______________________

**8. Was any azithromycin resistance detected in patients presenting with yaws?**

- *Yes*
- *No*
- *Unknown*

**9. Do you have any other information to share about molecular testing for azithromycin resistance?**

______________________________________________________________________

______________________________________________________________________

#### Section 3:  Molecular testing for syphilis

*These questions have been asked to assess what capacity you have for testing syphilis*

1. **Are there laboratories set up for molecular testing for syphilis?**
   - *Yes*
   - *No*
   - *Unknown*
2. **If yes, where are these laboratories located?**
   - *National Reference Laboratories*
   - *Regional labs*
   - *District labs*
   - *Sub-district (local)*
   - *Unknown*
3. **Is molecular testing for azithromycin-resistant syphilis available?**

*Available means an SOP in place in the lab, with trained staff and all reagents and equipment available to run these tests when requested*

- *Yes*
- *No*
- *Unknown*

4. **Is there an external quality assurance scheme in place for laboratories running molecular tests for either syphilis or yaws?**
   - *Yes*
   - *No*
   - *Unknown*

**5. Do you have any other information you would like to share with us about molecular diagnostics for syphilis?**

***______________________________________________________________________***

#### Section 4: Funding for molecular testing

1. **Is there sufficient funding available to pay for all sample collection equipment/consumables (e.g. swabs, tubes, labels) required for PCR? Sufficient means enough funding to cover the total costs**
   - *Yes*
   - *No*
   - *Unknown*
2. **What are the sources of funding for sample collection equipment?**

- *Health ministries*
- *Programme partner*
- *Other NGO*
- *WHO*
- *Other*
- *Unknown*

3. **Of those you have selected rank them in order of greatest contributor to funding. If not known move on to the next question.**

1.

2.

3.

4.

5.

6.

4. **Is there sufficient funding available to pay for PCR Reagents and consumables? Sufficient means enough funding to cover the total costs**
   - *Yes*
   - *No*
   - *Unknown*
5. **What are the sources of funding for PCR reagents and consumables?**
   - *Health ministries*
   - *Programme partner*
   - *Other NGO*
   - *WHO*
   - *Other*
   - *Unknown*
6. **Of those you have selected rank them in order of greatest contributor to funding. If not known move on to the next question.**

1.

2.

3.

4.

5.

6.

7. **Is there sufficient funding available to pay for sample shipment to the laboratory after collection of samples?**
   - *Yes*
   - *No*
   - *Unknown*
8. **Is there sufficient funding available to pay for staff costs to collect and analyse samples using PCR? Sufficient means enough funding to cover the total costs**
   - *Yes*
   - *No*
   - *Unknown*
9. **What are the sources of funding for staff costs?**
   - *Health ministries*
   - *Programme partner*
   - *Other NGO*
   - *WHO*
   - *Other*
   - *Unknown*
10. **Of those you have selected rank them in order of greatest contributor to funding. If not known move on to the next question.**

1.

2.

3.

4.

5.

6.

11. **Are there plans to set up additional laboratories for molecular testing?**
    - *Yes*
    - *No*
    - *Unknown*
12. **If q11 = yes, is there any funding available to set up these additional labs?**
    - *Yes*
    - *No*
    - *Unknown*
13. **If q12 = yes, where is this funding coming from?**
    - *Health ministries*
    - *Programme partner*
    - *Other NGO*
    - *WHO*
    - *Other*
    - *Unknown*

#### Section 5: Barriers to improving diagnostic capacity

*This final question is included as we wish to see what you believe are the biggest barriers/obstacles for increasing yaws diagnostic capacity. These barriers could be things like lack of funding, staff availability and/or difficulty accessing resources.*

1. **What barriers do you see to enhancing molecular diagnostic capacity?**
